# Three-month follow-up of heterologous vs homologous third vaccination in kidney transplant recipients

**DOI:** 10.1101/2022.02.22.22270838

**Authors:** Andreas Heinzel, Eva Schretzenmeier, Florina Regele, Karin Hu, Lukas Raab, Michael Eder, Christof Aigner, Rhea Jabbour, Constantin Aschauer, Ana-Luisa Stefanski, Thomas Dörner, Klemens Budde, Roman Reindl-Schwaighofer, Rainer Oberbauer

**Author notes:** Equally contributing first authors (Andreas Heinzel, Eva Schretzenmeier).

## Abstract

**Importance:** Response to SARS-CoV-2 vaccines in kidney transplant recipients (KTR) is severely reduced. Heterologous 3^rd^ vaccination combining mRNA and vector vaccines did not increase seroconversion at four weeks after vaccination but evolution of antibody levels beyond the first month remain unknown.

**Objective:** To assess changes in antibody response following a 3^rd^ vaccination with mRNA or vector vaccine in KTR from month one to month three after vaccination.

**Design, Setting and Participants:** Three-month follow-up (pre-specified secondary endpoint) of a single-center, single-blinded, 1:1 randomized, controlled trial on 3^rd^ vaccination against SARS-CoV-2 in 201 KTR who did not develop SARS-CoV-2 spike protein antibodies following two doses of an mRNA vaccine.

**Intervention(s):** mRNA (BNT162b2 or mRNA-1273) or vector (Ad26COVS1) as 3^rd^ SARS-CoV-2 vaccine

**Main Outcomes and Measures:** Main outcome was seroconversion at the second follow-up between 60-120 days after the 3^rd^ vaccination. Subsequently, higher cut-off levels associated with neutralizing capacity and protective immunity were applied (i.e. >15, >100, >141 and >264 BAU/mL). In addition, trajectories of antibody levels from month one to month three were analyzed. Finally, SARS-CoV-2 specific CD4 and CD8 T-cells at four weeks were compared among the 18 top responders in both groups.

**Results:** A total of 169 patients were available for the three-month follow-up. Overall, seroconversion at three months was similar between both groups (45% versus 50% for mRNA and vector group, respectively; OR=1.24, 95%CI=[0.65, 2.37], p=0.539). However, when applying higher cut-off levels, a significantly larger number of individual in the vector group reached antibody levels > 141 and > 264 BAU/mL at the three-month follow-up (141 BAU/mL: 4% vs. 15% OR=4.96, 95%CI=[1.29, 28.21], p=0.009 and 264 BAU/mL: 1% vs. 10% OR=8.75, 95%CI=[1.13, 396.17], p=0.018 for mRNA vs. vector vaccine group, respectively). In line, antibody levels in seroconverted patients further increased from month one to month three in the vector group while remaining unchanged in the mRNA group (median increase: mRNA= 1.35 U/mL and vector = 27.6 U/mL, p = 0.004). Of particular note, there was no difference in the CD4 and CD8 T-cell response between the mRNA and vector vaccine group at month one.

**Conclusions and Relevance:** Despite a similar overall seroconversion rate at three months following 3^rd^ vaccination in KTR, a heterologous 3rd booster vaccination with Ad26COVS1 resulted in significantly higher antibody levels in responders.

**Trial Registration:** EurdraCT: 2021-002927-39

## Introduction

Vaccine response in kidney transplant recipients (KTR) is severely reduced due to the mandatory immunosuppressive medication following transplantation. Subsequently, a significant number of KTR remains at risk for SARS-CoV-2 infection despite vaccination.^1,2^ Strategies to improve vaccine response in this high-risk vulnerable population for severe COVID-19 are urgently needed.

We recently conducted a randomized single-blinded controlled trial in 200 patients comparing a homologous vs heterologous vaccination strategy in KTR who did not develop SARS-CoV-2 spike protein-specific antibodies after two doses of an mRNA vaccine: Overall, 39% of patients developed antibodies at four weeks after the 3^rd^ dose, with no statistically significant difference between an additional dose of the same mRNA vaccine as used for the initial prime/boost vaccination (BNT162b2 or mRNA-1273, 35% response rate) or a vector vaccine (Ad26COVS1, 42% response rate).^3^

Other recent reports, however, suggest a more pronounced induction of both, a SARS-CoV-2 specific CD4 T cell response and antibodies following heterologous vaccination that includes a vector-based vaccine in transplant recipients.^4^ In line, heterologous 3^rd^ vaccination also increased overall T-cell response in patients treated with B-cell depleting therapy.^5^

Most analysis to date were limited to observation within the first four weeks after 3^rd^ vaccination. Another recent observational study from France reported changes in antibody levels in KTR form one month to three months after a 3^rd^ mRNA vaccine showing a significant reduction in antibody levels.^6^ However, data on trajectories of antibody levels beyond the first month following heterologous vaccination remain unknown. In the current analysis of our randomized controlled trial (RCT) including follow-up data until month three, we aimed to assess changes in antibody over time (months one to month three) following homologous vs heterologous 3^rd^ vaccination.

## Methods

### Study cohort and trial design

Study participants were followed-up for antibody assessment at the outpatient’s transplant clinic of the Medical University of Vienna for a second follow-up (FU) between 60-120 days after the 3^rd^ vaccine dose (*three months FU*, pre-specified secondary endpoint). Details on randomization and treatment have been reported before.^3^ In short: 200 patients without detectable SARS-CoV-2 specific antibodies following two doses of an mRNA vaccine were randomized to a 3^rd^ dose of the same mRNA vaccine (mRNA group) or a dose of the vector vaccine Ad26COVS1. Clinical endpoints (death, COVID-19) were recorded for all study participants throughout the observation period.

### Assessment of the humoral response

Antibody response was evaluated using the Roche Elecsys anti–SARS-CoV-2 S enzyme immunoassay (Roche, Switzerland) detecting antibodies against the receptor-binding domain of the SARS-CoV-2 spike protein (the cutoff at 0.8 U/mL according to the manufacturer’s instructions). As additional endpoints, we applied higher cut off levels that were also reported as secondary endpoints at the one-month FU and that are associated with neutralizing capacity as well as reduced risk for COVID-19 infection: >100 U/mL^7^, >141 BAU/mL^8^ and >264 BAU/mL.^9^ BAU/mL were converted to U/mL based on the conversion formula: U/mL=0.972*BAU/mL.

### Assessment of T-cell response

Besides the humoral response we further analyzed SARS-CoV-2-specific CD4 and CD8 T-cell responses among humoral top responders at four weeks in both groups (n=18 per group). The T-cell stimulation flow cytometric (FC) assessment of SARS-CoV-2 specific T-cells have been described before.^10,11^ In brief, peripheral blood mononuclear cells (PBMC) were isolated by Ficoll-Paque density gradient centrifugation and cryopreserved until further analysis. For the identification of SARS-CoV-2 specific T-cells 3-5 × 10^6^ PBMCs were incubated for 18h with overlapping 15-mer peptides covering the complete SARS-CoV-2 spike protein wild type variant (1 ug/ml per peptide; JPT, Germany) and subsequently subjects to FC analysis. SARS-CoV-2 specific CD4 T-cells were identified based on CD154 and CD137 co-expression whereas co-expression of CD137 and IFN-γ was used for CD8 T-cells. Patients were considered having SARS-CoV-2 specific T-cells when the number of identified cells in the stimulated sample exceeded the number such cells in the unstimulated sample by at least twofold. To account for patient specific background activation frequencies of activated cells detected in control samples were subtracted from the stimulated samples prior to any subsequent analysis.

### Statistical analysis

Patient demographics for continuous variables were reported as median and interquartile range except for patient age which was reported as mean and standard deviation. Categorical variables were described by frequency and percentage. Differences between treatment groups for continuous and categorical variables were assessed by Wilcoxon rank sum test and Fisher’s exact test, respectively. Wilcoxon rank sum tests were used for all comparison of absolute antibody concentrations as well as antibody level differences from one-month to three-month FU and detectable fractions of SARS-CoV-2 specific T-cells between groups. The number of seroconverted patients, number of patients with SARS-CoV-2 specific T-cells and the number of patients exceeding defined antibody level cutoffs between groups were evaluated by means of Fisher’s exact test.

## Results

### Study population

From the initially enrolled n=201 patients, blood samples from 169 patients were available for the three month FU analysis of vaccine efficacy: 85 and 84 patients in the mRNA and vector group, respectively (CONSORT Flow Chart is provided in **Figure 1**). Patient characteristics are provided in **Table 1**. There was no statistically significant difference between the mRNA and vector vaccine groups. Overall, eight deaths and seven SARS-CoV-2 infections occurred in the study population within the observation period (death: four vs four; COVID-19: three vs four for mRNA vs vector vaccine group, respectively). All COVID-19 cases occurred in vaccine no-/low-responders; three patients had severe COVID-19 requiring ICU treatment (two patients in the vector groups died as well as one patient from mRNA group who was on extra-corporal membrane oxygenation).

**Table 1:** Demographics of the study population

| Variable | mRNA | vector |
| --- | --- | --- |
| N | 85 | 84 |
| Mean (SD) age, y | 61 (13) | 61 (12) |
| Sex |  |  |
| Female | 37 (44) | 34 (40) |
| Male | 48 (56) | 50 (60) |
| Time since KTX, y | 4.8 [2.4-8.6] | 4.9 [1.6-7.4] |
| No. of KTX |  |  |
| 1 | 64 (75) | 66 (79) |
| 2 | 15 (18) | 13 (15) |
| 3 | 4 (5) | 4 (5) |
| 4 | 2 (2) | 0 (0) |
| 5 | 0 (0) | 1 (1) |
| Donor type (living) | 14 (16) | 18 (21) |
| Initial vaccinations (mRNA-1273) | 27 (32) | 27 (32) |
| Maintenance immunosuppression |  |  |
| Belatacept, MMF, steroids | 6 (7) | 6 (7) |
| Belatacept, azathioprine, steroids | 0 (0) | 1 (1) |
| Cyclosporin A, MMF, steroids | 1 (1) | 4 (5) |
| Cyclosporin A, MMF | 3 (4) | 1 (1) |
| Cyclosporin A, azathioprine, steroids | 1 (1) | 0 (0) |
| MMF, steroids | 1 (1) | 1 (1) |
| Tracolumus, MMF, steroids | 66 (78) | 62 (74) |
| Tracolumus, MMF | 1 (1) | 3 (4) |
| Tracolumus, azathioprine, steroids | 4 (5) | 3 (4) |
| Tracolumus, steroids | 2 (2) | 2 (2) |
| Tracolumus, leflunomide, steroids | 0 (0) | 1 (1) |
| ATG in past year | 1 (1) | 2 (2) |
| Nontriple immunosuppression | 7 (8) | 7 (8) |
| Time between second and third vaccination, d | 78 [55-87] | 80.5 [57-90.25] |
| Time between third vaccination and one-month follow-up visit, d | 31 [28-32] | 30 [28-33] |
| Time between third vaccination and three-month follow-up visit, d | 81 [74-88] | 76 [69-89] |

**Figure 1:**
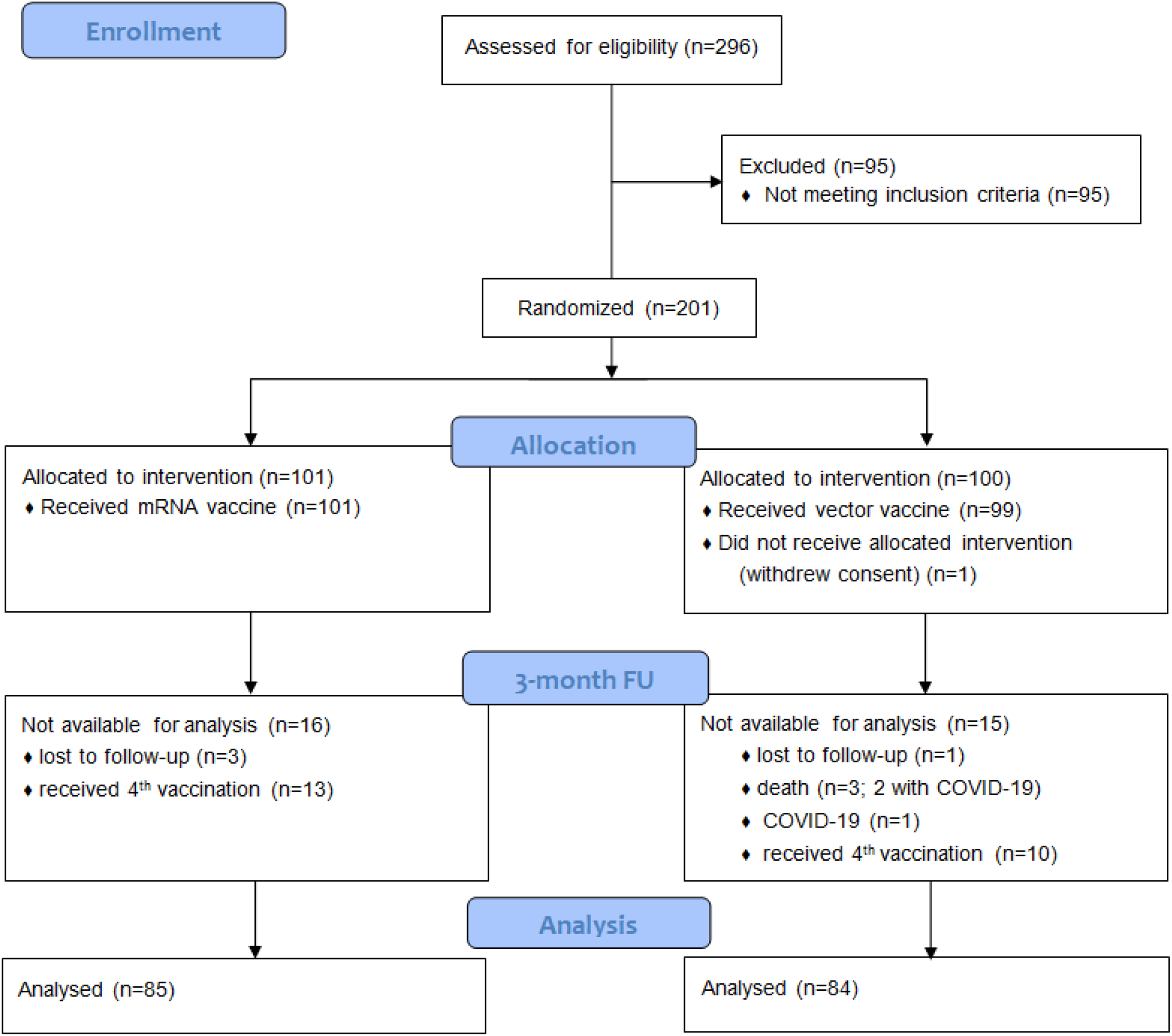
CONSORT Flow Chart for the 3 moth Follow-up. Blood samples for evaluation of vaccine efficacy at the three-month FU were available for 169 of the initially enrolled 201 patients: One patient withdrew consent before vaccination, 23 patients were excluded after they had received a 4^th^ vaccine dose before completing the three-month FU visit; one patient died following myocardial infarction, two patients died due to COVID-19, one patient had mild COVID-19 and four patients had no blood-draw within the observation period.

### Humoral immune response

Overall response rate to the 3^rd^ vaccine dose at the three-month FU was 47% with no statistically significant difference in seroconversion between the mRNA and vector vaccine group (mRNA: 45% and vector: 50% OR=1.24, 95%CI=[0.65, 2.37], p=0.539). Absolute antibody titers between the two groups were also not significantly different (median mRNA: 0.2 U/mL and vector: 0.81 U/mL, p=0.104). However, when examining higher antibody cut-off levels that were also included in our primary analysis at the one-month FU, we observed that a significantly higher number of patients in the vector group reached antibody levels above 141 and 264 BAU/mL (141 BAU/mL: 4% vs. 15% OR=4.96, 95%CI=[1.29, 28.21], p=0.009 and 264 BAU/mL: 1% vs. 10% OR=8.75, 95%CI=[1.13, 396.17], p=0.018, for mRNA vs. vector vaccine group, respectively, **Table 2**). In contrast, no difference between the groups was observed for any of the antibody level cut-offs at the one-month FU (**Table 2**).

**Table 2:**
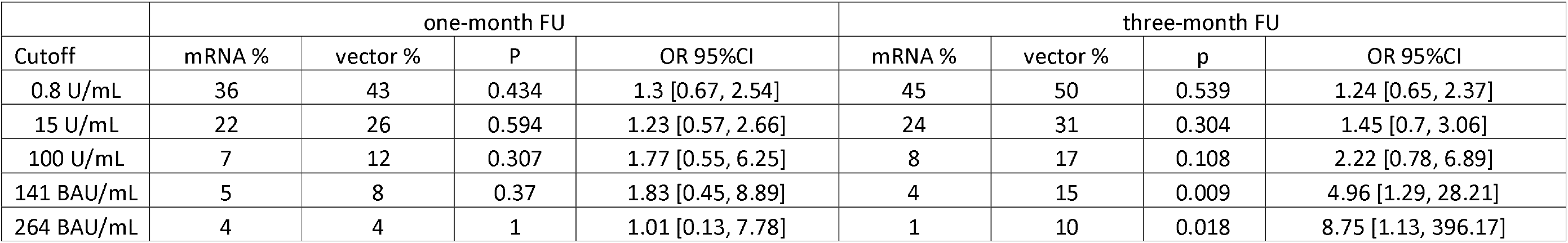
Response rate to 3^rd^ SARS-CoV-2 vaccination at different pre-specified cut-off levels for the one- and three-month follow-up

### Change in serostatus between month one versus month three

In both groups a comparable number of patients who had not seroconverted at the one-month FU became seropositive in the subsequent months (8% and 8% OR=1.01, 95%CI=[0.29, 3.56], p=1 for mRNA and vector, respectively). With the exception of a single patient in the vector group all patients who showed seroconversion at the one-month FU had antibody levels above the 0.8 U/mL cutoff at the three-month FU. **Figure 2A** visualizes changes in serostatus including increase above 141 BAU/mL as surrogate for protective immunity.

**Figure 2:**
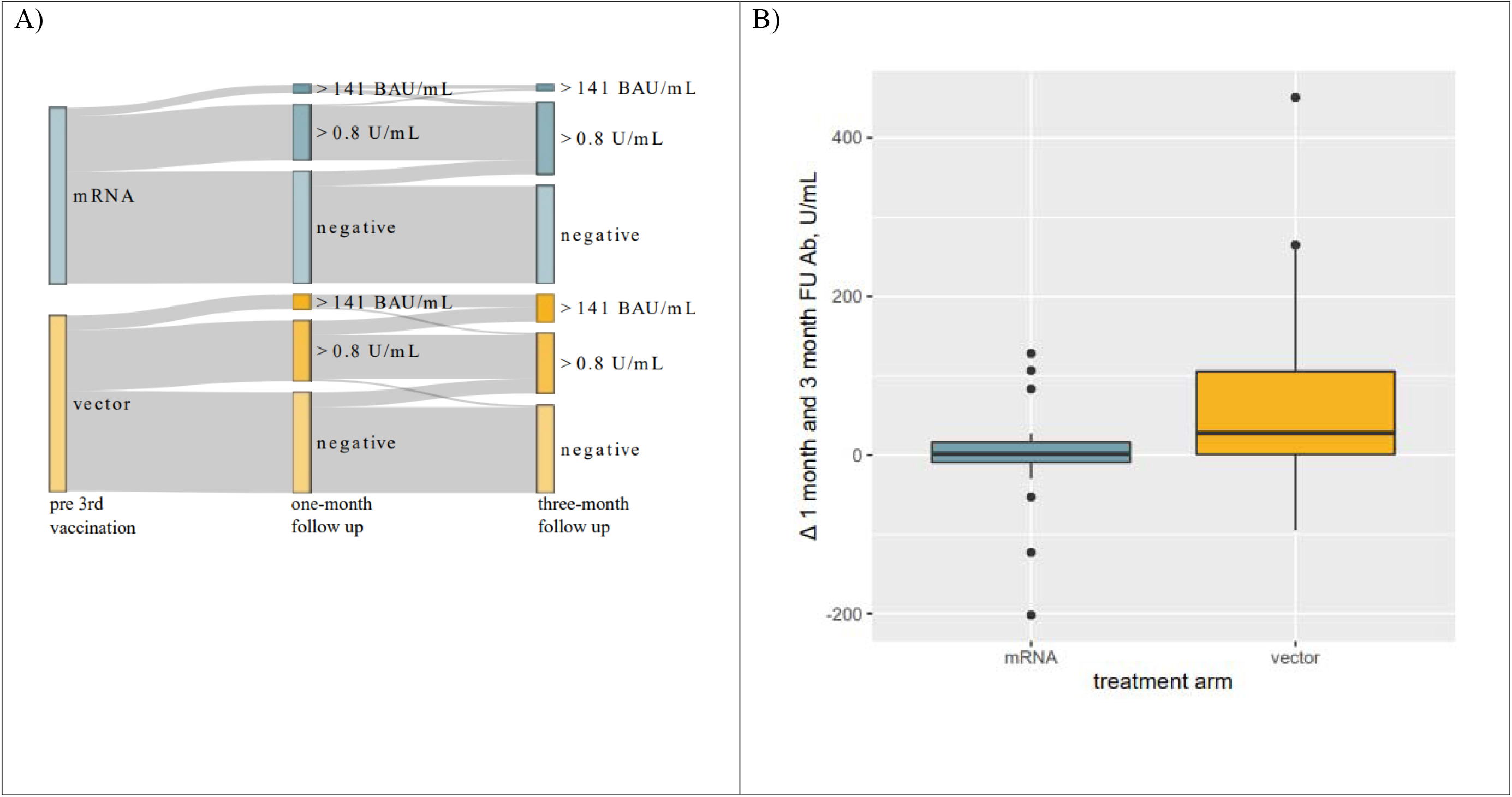
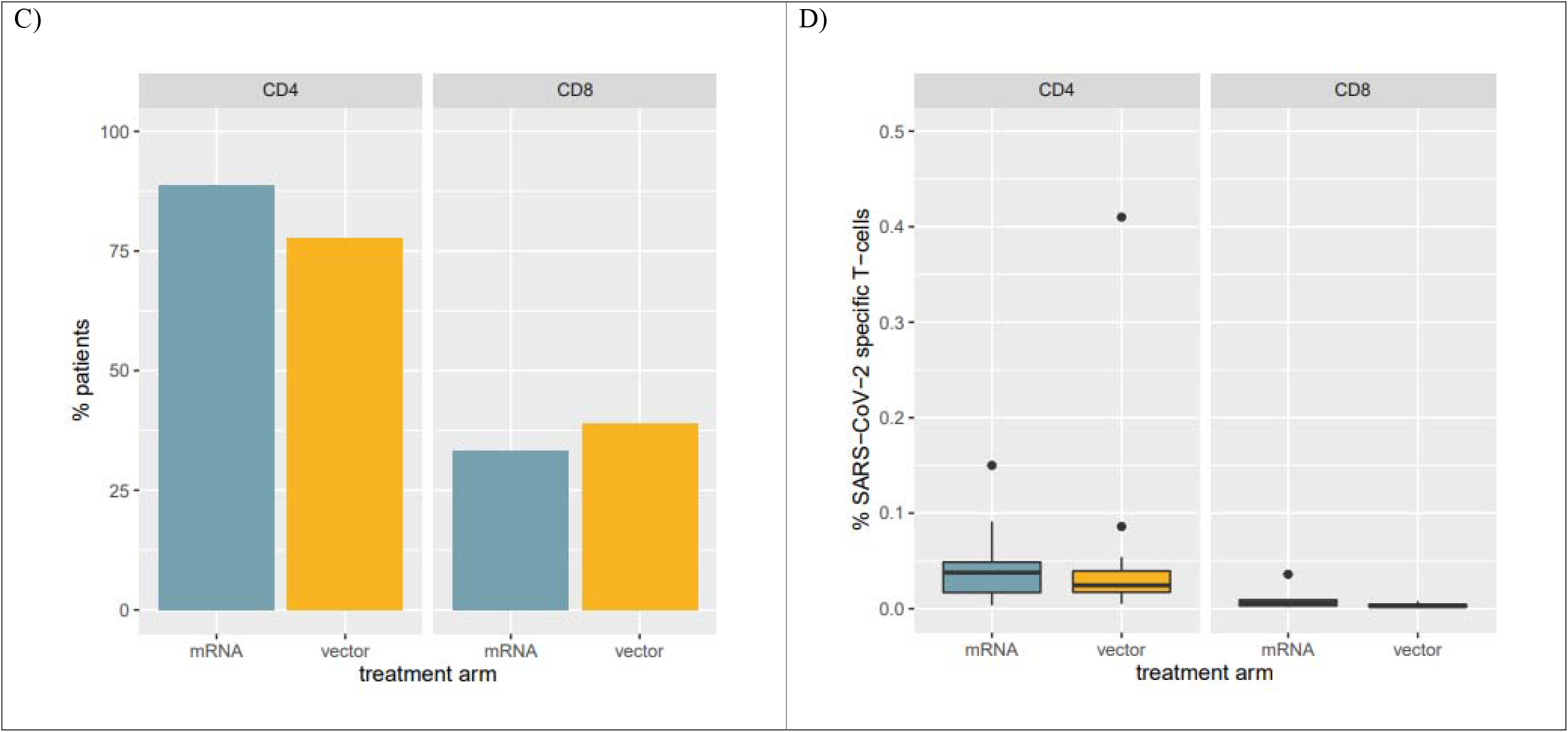
Response to vaccination. Panel A: Sankey Diagram visualizing changes in response rate to 3^rd^ vaccination. A significantly larger proportion of individuals developed antibody levels > 141 BAU/mL. Panel B: Boxplots visualizing changes in antibody levels from one-to three-month FU in patients who seroconverted within one-month after receiving their 3^rd^ vaccination. Antibody levels in individuals receiving a heterologous 3^rd^ vaccination further increased while remaining unaltered in patients receiving mRNA vaccines. Panel C: Percentage of patients with SARS-CoV-2 specific CD4 and CD8 T-cells among the top humoral responders at the one-month FU. Panel D: Percentages of SARS-CoV-2 specific T-cells in patients with SARS-CoV-2 specific CD4 and CD8 T-cells.

### Evolution of antibody levels beyond the first month

Of particular note, evolution of antibody levels in patients with seroconversion at the one-month FU differed significantly between the two groups. Antibody levels in the vector group further increased after the one-month FU while remaining approximately unchanged in the mRNA group (median of differences mRNA: 1.35 U/mL and vector: 27.6 U/mL, p=0.004, **Figure 2B**). Consequently, absolute antibody levels were significantly different between the two treatment groups at the three-month FUP (median mRNA: 25.8 U/mL and vector: 77.7 U/mL, p = 0.038), even though, they were not significantly different at the one-month FU (mRNA: 19.7 U/mL and vector: 22.1 U/mL, p = 0.753).

### T-cell response

We also analyzed the T-cell responses at the one-month FU1 in 18 patients among the top responders to the 3^rd^ vaccine from both groups to see if the subsequent increase in antibody levels in the vector group was preceded by a higher SARS-CoV-2 specific T-cell response. After the 3^rd^ vaccination 83% and 36% of patients had SARS-CoV-2 specific CD4 and CD8 cells, respectively. The number of patients with SARS-CoV-2 specific CD4 and CD8 T-cells was comparable between the treatment groups (CD4 mRNA: 89% and vector: 78% OR=0.45, 95%CI=[0.04, 3.68], p=0.658; CD8 mRNA: 33% and vector: 39% OR=1.26, 95%CI=[0.27-6.19], p=1, Figure 2C). In patients with SARS-CoV-2 specific CD4 and CD8 T-cells a median of 0.033% and 0.003% overall CD4 and CD8 cells were SARS-CoV-2 specific. Interestingly, these numbers were also comparable between the two treatment groups (CD4 mRNA: 0.038% and vector: 0.024% p=0.547; CD8 mRNA: 0.006% and vector: 0.003% p=0.295, Figure 2D).

## Discussion

In this three-month FU analysis of our RCT on homologous vs heterologous 3^rd^ vaccination in KTR we observed an increase in antibody levels from month one to month three in individuals receiving a heterologous 3^rd^ vaccination dose with the vector vaccine Ad26COVS1. In contrast, antibody levels in individuals receiving a homologous 3^rd^ vaccination with an additional dose of an mRNA remained unchanged from the one-month FUP to the three-month FUP resulting in overall lower antibody levels in the homologous vaccination group. Consequently, there was a significantly higher number of individuals with antibody levels above predefined antibody thresholds associated with neutralizing capacity despite a comparable overall seroconversion rate. Especially in the face of new variants that evade immune response, higher antibody levels providing broader coverage are needed for infection prevention but cut-off levels conveying protective immunity remain undefined.^12^ Clinical endpoints were similar between both intervention groups.

Interestingly, there was no difference in the SARS-CoV-2 specific CD4 or CD8 T-cell response at four weeks after vaccination comparing homologous or heterologous vaccination strategies. This contrasts with other reports suggesting higher levels of T-cell response following heterologous vaccination^5,13^, although clear thresholds or correlates of T cell protection remain to be delineated. However, other studies used the vector vaccine ChAdOx1 as opposed to Ad26COVS1. Impact of heterologous vaccination on antibody levels in these studies was inconclusive with one suggesting higher antibody levels in the heterologous group (KTR) while another showed a lower seroconversion rate in the heterologous vaccination group (patients treated with rituximab).

Despite similar overall seroconversion rates and comparable antibody levels at four weeks, heterologous 3^rd^ boost vaccination using Ad26COVS1 results in significantly higher antibody levels but not CD4 or CD8 responses in KTR over a three-month follow-up period compared to additional homologous vaccination.

## Data Availability

All data produced in the present study are available upon reasonable request to the authors

